# Opportunistically Detecting Signs of Hypertension on a Consumer Smartwatch

**DOI:** 10.64898/2025.12.10.25341972

**Authors:** Paolo Di Achille, Lawrence Cai, Jiang Wu, Mingwu Gao, Bhavna Daryani, Jonathan Wang, Utkarsh Khanna, Ho Ko, Anupam Pathak, Mark Malhotra, Shwetak Patel, Jacqueline B. Shreibati, Stephen P. Juraschek, Pramod Rudrapatna, Matthew Thompson, Ming-Zher Poh

## Abstract

Hypertension is a silent killer, with over half of affected adults unaware of their condition^1,2^. This lack of awareness is a major concern, as early intervention is critical for preventing major adverse cardiovascular events^3,4^. While cuffless wearable blood pressure (BP) monitors offer comfort and convenience, their reliance on periodic calibration and inconsistent accuracy have limited their clinical adoption^5,6^. Here we show that applying artificial intelligence (AI) pre-trained on almost 500,000 hours of data to multimodal waveforms (photoplethysmography and accelerometry) recorded using a widely available consumer smartwatch (AI-PPG-ACC-HTN), without any cuff calibration, can detect hypertension with accuracy levels comparable to traditional cuffed BP devices in the existing clinical framework, including both initial and confirmatory screening. We validated AI-PPG-ACC-HTN in a prospective, multicenter study of 196 diverse participants free from known cardiovascular disease and antihypertensive medication against gold-standard 24-hour ambulatory BP monitoring. Over seven days of real-world monitoring, AI-PPG-ACC-HTN detected hypertension with a sensitivity of 65.8% (95% CI, 54.0%-76.3%), specificity of 90.0% (83.2-94.7), and positive predictive value (PPV) of 80.6% (68.6-89.6). In comparison, initial office BP screening achieved a sensitivity of 55.3% (43.4-66.7), specificity of 90.0% (83.2-94.7) and PPV of 77.8% (64.4-88.0). For confirmatory testing of participants with elevated BP identified by initial office BP screening (N=48), AI-PPG-ACC-HTN detected hypertension with a sensitivity of 78.4% (61.8-90.2), specificity of 90.9% (58.7-99.8) and PPV of 96.7% (82.8-99.9). Comparatively, repeat office BP achieved a sensitivity of 67.6% (50.2-82.0), specificity of 63.6% (30.8-89.1) and PPV of 86.2% (68.3-96.1); multiday home BP monitoring achieved a sensitivity of 89.2% (74.6-97.0), specificity of 81.8% (48.2-97.7) and PPV of 94.3% (80.8-99.3). These results highlight an opportunity for consumer smartwatches to facilitate population-level opportunistic hypertension screening, offering an accessible tool to address this major public health challenge.

## Introduction

Hypertension is the top modifiable risk factor for cardiovascular disease and all-cause mortality worldwide^7^, yet globally around half of adults with hypertension are unaware that they have the condition. Most individuals with the condition have no signs or symptoms and their hypertension is typically only discovered through incidental screening^8,9^. Despite the recommendation by the US Preventive Services Task Force (USPSTF) to screen adults annually for high blood pressure (BP)^10^, there remains a concerning gap in awareness of hypertension in the general population that represents a major public health challenge. A recent study examining hypertension control revealed that more than one-half (57.8%) of US adults were unaware they had uncontrolled hypertension, even among those reporting 2 or more health care visits in the past year (51.8%)^2^. Lack of awareness was notably high at 68.4% among those aged 18 to 44 years. Other barriers to detecting hypertension can include limited access to healthcare, as well as varying clinician practices towards screening^11^, all of which can further contribute to suboptimal management of cardiovascular health and contribute to major adverse events later in life^3,4^.

The pressing need to improve hypertension awareness may be meaningfully addressed by leveraging the increasingly widespread adoption of consumer wearable devices capable of performing photoplethysmography (PPG). While the potential for PPG to reflect cardiovascular health has been known for decades^12,13^, translating this into reliable, calibration-free BP monitoring has proven challenging^5,6^. A major concern is that these devices typically require individual user calibration with cuff BP measurements, meaning they primarily track BP changes relative to calibration rather than measuring absolute BP^6^. Recent research leveraging deep learning has made progress with studies demonstrating the potential for calibration-free hypertension detection, as a binary classifier, using PPG signals^14,15^, culminating in the notable launch of a PPG-based smartwatch hypertension screening feature cleared by the US Food and Drug Administration (FDA)^16^. While some of these approaches rely on the accelerometer (ACC) to identify periods of stillness or when a user is seated, they do not leverage information in the ACC signals themselves that may be associated with BP^17^. Importantly, a common limitation of studies in support of these features is the use of home blood pressure monitoring as the reference standard rather than 24-hour ambulatory blood pressure monitoring (ABPM), which may obscure their true performance. Furthermore, the performance of PPG-based consumer wearables against the established clinical framework for hypertension screening^10,18^ is a critical evidence gap for clinical translation.

In this work, we develop and validate a multimodal artificial intelligence (AI) system for detecting hypertension (AI-PPG-ACC-HTN) using concomitant PPG and ACC signals acquired from a consumer smartwatch and a user’s height as a proxy for arterial path length. We compare its performance with traditional cuffed BP devices in the current hypertension screening framework against 24-hour ABPM in a multisite prospective clinical study of 196 participants in free-living conditions. We demonstrate that AI-PPG-ACC-HTN detects hypertension with an accuracy comparable to current US clinical practice of initial screening using office-based BP measurements and confirmatory home-based BP measurements, highlighting an opportunity for consumer smartwatches to facilitate population-level opportunistic hypertension screening and address this major public health challenge.

## Results

### Overview of system

Our AI system leverages multimodal information contained in the wrist PPG signal and ACC (Fig. 1A). The ACC signals provide information on the magnitude of wrist motion associated with each pulse and, during times of low motion, may also capture cardiac timing information in the form of the ballistocardiogram (BCG)^19,20^. Pulse wave analysis of the PPG contour and pulse transit time between the BCG and PPG signal have been separately associated with BP and arterial stiffness. Informed by the physiological underpinnings of these input signals, our system is innovative in taking an AI-based multimodal approach to jointly encode the PPG and ACC signals. Instead of defining hand-engineered features, we trained a 1-D deep convolutional neural network (Waveform Encoder) in a self-supervised manner to create rich embeddings of 256 dimensions from inputs of 15-second PPG and corresponding accelerometer signals without the need for hypertension or BP labels. This was undertaken using pooled unlabeled smartwatch data collected from 2,109 participants across a series of studies conducted in free-living conditions (Supplemental Table 1); these data amounted to 492,347 hours. To confirm that this pre-training methodology learns a physiologically relevant embedding, we conducted a separate, exploratory analysis on a large external cohort (UK Biobank, N=76,268) using a refitted, unimodal PPG-only version (UK Biobank did not have concomitant ACC data) of the encoder (Supplemental Information). Next, we mean-aggregated the embeddings across the entire duration of wear to produce a single embedding for each person. Lastly, we trained a logistic regression classifier to detect hypertension from the person-level embeddings with the individual’s height as an additional input to approximate the arterial path length^21^ using smartwatch data collected from a diverse population of 663 participants from 44 different states across the US who also performed 24-hour ABPM to provide reference labels for hypertension (Table 1). We found that adding ACC and height improved performance during development (Supplemental Fig. 1, Supplemental Table 2).

**Figure 1.**
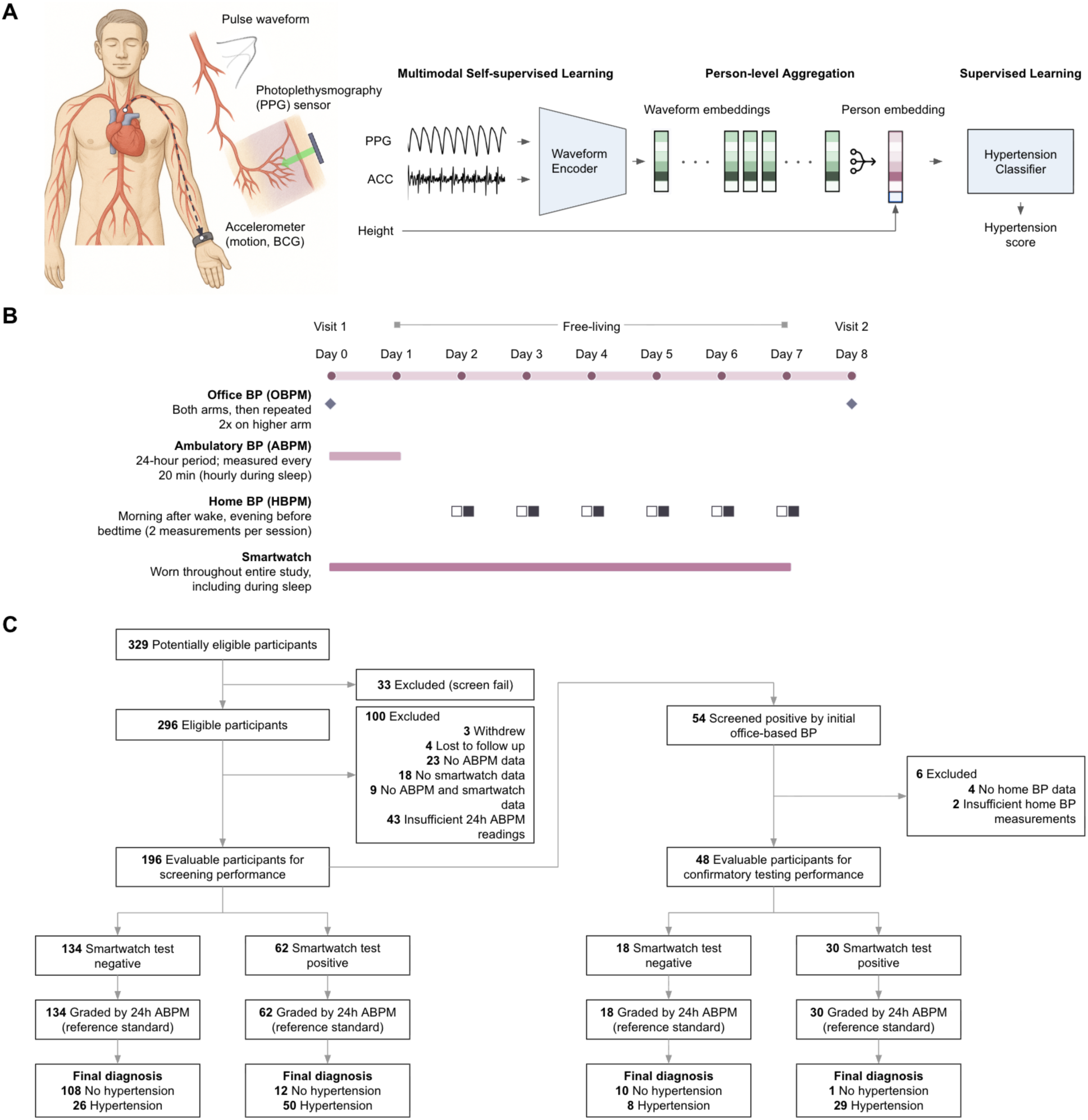
System overview and validation of a smartwatch-based AI system for detecting hypertension (AI-PPG-ACC-HTN). (A) The AI system uses photoplethysmography (PPG) and accelerometer (ACC) signals recorded using a consumer smartwatch to create waveform embeddings. The waveform embeddings are aggregated over time (days) into a person-level embedding, which, along with a user’s height, are used to predict their hypertension status. (B) AI-PPG-ACC-HTN was externally validated in a prospective clinical validation study. (C) Flow of participants through the validation study. BCG: ballistocardiogram.

**Table 1.**
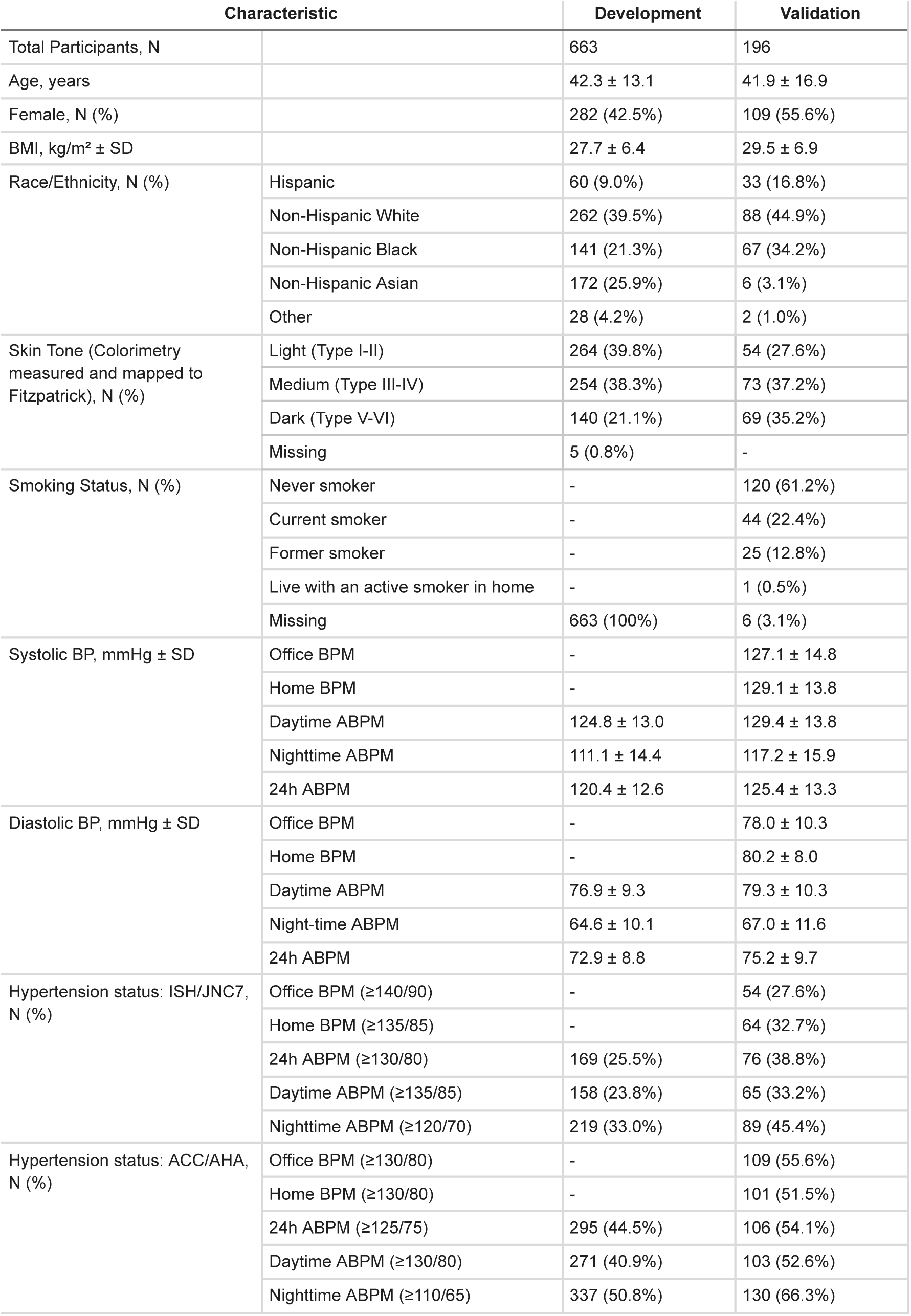
Participant Characteristics.

### Prospective validation study

To evaluate the performance of our AI system, we conducted a prospective clinical study across five sites in the US. The study population comprised individuals aged 18 years or older who did not have a history of known cardiovascular disease (previous myocardial infarction, stroke, cardiac arrest, ischemic heart disease, atrial fibrillation, heart failure, or aortic valve disorder) and were not taking antihypertensive medication. A total of 68.4% of participants had a previous elevated office BP reading with no hypertension diagnosis or a diagnosis of hypertension but were not prescribed BP-lowering medications.

This study involved measurements of smartwatch data, office BP, home BP, and reference ABPM to allow comparisons between traditional cuff-based BP and the proposed approach (Fig. 1B). Enrolled participants attended two study visits and completed an at-home, free-living study over 7 consecutive days. Initial office-based BP measurements (office BPM) were performed following international guidelines^22^, measuring both arms and adopting the higher arm for two further readings used for subsequent analysis, using an automated oscillometric device with the appropriate BP cuff size based on measured arm circumference. Participants were then outfitted with an FDA-cleared ABPM, provided with an automated home BP monitor, and trained on the proper use of each device. Starting from the end of the first study visit, participants were instructed to wear the ABPM for 24 hours, and the smartwatch on the opposite wrist throughout the duration of the free-living study. On the 2nd through 7th days, participants performed two sets of daily home BP measurements (home BPM) following the guidelines: one set in the morning after rising, and one set in evening before bedtime. Each set of readings consisted of two BP measurements separated by approximately 2 minutes in between. During the second (final, on 8th day) study visit, two further office-based BP measurements were performed. Full study details are provided in the Methods section.

We enrolled 296 participants, of which 196 (66.2% completion rate) had sufficient data (with complete office BPM and ABPM data), and were included in our analysis (Fig. 1C). The major reasons for exclusion from analysis were missing or incomplete ABPM data (75/100 of excluded participants), especially the lack of ABPM measurements during sleep. Comparing baseline characteristics of the full enrolled cohort versus the final analysis cohort (Supplemental Table 3), we found no statistically significant difference in age, sex, BMI or office BPM systolic BP (SBP) and diastolic BP (DBP). In this final analysis cohort (Table 1), the mean ± SD age was 41.9 ± 16.9 years, 55.6% were female, 34.2% were non-Hispanic Black, and 16.8% were Hispanic. A wide range of skin tones were represented as objectively quantified by individual typology angle (ITA°) based on colorimetry ranging from -71.5° to 88.8°. Grouping by three skin tone cohorts yielded 27.6% in light, 37.2% in medium and 35.2% in dark skin tones groups mirroring the Fitzpatrick skin types of I-II, III-IV and V-VI.

The mean ± SD of the initial office BPM SBP / DBP was 127.1 ± 14.8 / 78.0 ± 10.3 mmHg. The mean ± SD number of ABPM measurements was 51.9 ± 13.3; 33.0 ± 9.8 measurements were taken during daytime and 5.0 ± 1.3 measurements during nighttime. Based on ABPM, the mean ± SD 24-hour SBP / DBP was 125.4 ± 13.3 / 75.2 ± 9.7 mmHg; daytime and nighttime SBP / DBP were 129.4 ± 13.8 / 79.3 ± 10.3 mmHg and 117.2 ± 15.9 mmHg / 67.0 ± 11.6 mmHg, respectively. The mean ± SD number of days with home BPM measurements was 6.0 ± 1.4. The mean ± SD of home BPM SBP / DBP was 129.1 ± 13.6 / 80.2 ± 8.0 mmHg. Participants wore the smartwatch for a mean ± SD of 7.3 ± 1.3 days, with an average of 14.7 ± 3.5 hours per day.

### AI System detects hypertension with comparable performance to office and home BP

We used 24-hour ABPM, considered the clinical gold standard for the correct diagnosis of hypertension, as the reference standard. Hypertension was defined as mean 24-hour SBP/DBP ≥130/80 mmHg following international guidelines^22^. The corresponding criteria for hypertension classification for office BPM was SBP ≥140 mmHg or DBP ≥90 mmHg; for home BPM it was SBP ≥135 mmHg or DBP ≥85 mmHg^18^.

In this study population, 38.8% of participants tested positive for hypertension based on 24-hour ABPM. Our AI system achieved moderate sensitivity (65.8% [95% CI, 54.0%-76.3%]), high specificity (90.0% [95% CI, 83.2%-94.7%]), high PPV (80.6% [95% CI, 68.6%-89.6%]), moderate positive likelihood ratio (6.6 [95% CI, 3.8-11.5]) and small negative likelihood ratio (0.4 [95% CI, 0.3-0.5]) for detecting hypertension (Table 2). In comparison, a logistic regression model using demographic data (age, sex, BMI) alone yielded poor sensitivity (34.2% [95% CI, 23.7%-46.0%]), high specificity (90.0% [95% CI, 83.2%-94.7%]), moderate PPV (68.4% [95% CI, 51.3%-82.5%]), small positive likelihood ratio (LR+, 3.4 [95% CI, 1.8-6.4]) and poor negative likelihood ratio (LR-, 0.7 [95% CI, 0.6-0.9]) for detecting hypertension. Initial screening office BPM had low sensitivity (55.3% [95% CI, 43.4%-66.7%]), high specificity (90.0% [95% CI, 83.2%-94.7%]), moderate PPV (77.8% [95% CI, 64.4%-88.0%]), moderate LR+ (5.5 [95% CI, 3.1-9.8]) and small LR- (0.5 [95% CI, 0.4-0.6]) for detecting hypertension.

**Table 2.** Test Accuracy of a Demographics-based Model, Screening Office Blood Pressure Monitoring (≥140/90 mmHg), and AI-PPG-ACC-HTN to Identify Hypertension Based on 24-hour Ambulatory Blood Pressure Monitoring (≥130/80 mmHg)

| **Model, N=196** | **Sensitivity** | **Specificity** | **PPV** | **NPV** | **LR+** | **LR-** | **AUROC** |
| --- | --- | --- | --- | --- | --- | --- | --- |
| Demographics | 34.2<br>(23.7, 46.0) | 90.0<br>(83.2, 94.7) | 68.4<br>(51.3, 82.5) | 68.4<br>(60.5, 75.5) | 3.4<br>(1.8, 6.4) | 0.7<br>(0.6, 0.9) | 0.775<br>(0.711, 0.837) |
| Office BPM (Visit 1) | 55.3<br>(43.4, 66.7) | 90.0<br>(83.2, 94.7) | 77.8<br>(64.4, 88.0) | 76.1<br>(68.2, 82.8) | 5.5<br>(3.1, 9.8) | 0.5<br>(0.4, 0.6) | 0.865<br>(0.814, 0.913) |
| Smartwatch<br>(AI-PPG-ACC-HTN) | 65.8<br>(54.0, 76.3) | 90.0<br>(83.2, 94.7) | 80.6<br>(68.6, 89.6) | 80.6<br>(72.9, 86.9) | 6.6<br>(3.8, 11.5) | 0.4<br>(0.3, 0.5) | 0.856<br>(0.798, 0.908) |

For confirmatory testing of participants with elevated BP identified by initial office BP screening (Table 3), AI-PPG-ACC-HTN detected hypertension with a high sensitivity of 78.4% (95% CI, 61.8%-90.2%), high specificity of 90.9% (95% CI, 58.7%-99.8%), high PPV of 96.7% (95% CI, 82.8%-99.9%), moderate LR+ of 8.6 (95% CI, 1.3-56.3) and moderate LR- of 0.2 (95% CI, 0.1-0.5). In comparison, repeat office BP achieved a moderate sensitivity of 67.6% (95% CI, 50.2%-82.0%), moderate specificity of 63.6% (95% CI, 30.8%-89.1%), high PPV of 86.2% (95% CI, 68.3%-96.1%), poor LR+ of 1.9 (95% CI, 0.8-4.2) and poor LR- of 0.5 (95% CI, 0.3-1.0); home BP monitoring over ≥ 3 days achieved a high sensitivity of 89.2% (95% CI, 74.6%-97.0%), high specificity of 81.8% (95% CI, 48.2%-97.7%), high PPV of 94.3% (95% CI, 80.8%-99.3%), small LR+ of 4.9 (95% CI, 1.4-17.3) and moderate LR- of 0.1 (95% CI, 0.1-0.3). These results indicate that the AI-PPG-ACC-HTN system detected hypertension with an accuracy comparable to cuff-based BP devices for both screening and confirmatory testing purposes.

**Table 3.** Test Accuracy of a Confirmatory Office Blood Pressure Monitoring (≥140/90 mmHg), Home Blood Pressure Monitoring (≥135/85 mmHg), and AI-PPG-ACC-HTN to Identify Hypertension Based on 24-hour Ambulatory Blood Pressure Monitoring (≥130/80 mmHg) in Participants Who Initially Screened Positive.

| Model, N=48 | Sensitivity | Specificity | PPV | NPV | LR+ | LR- | AUROC |
| --- | --- | --- | --- | --- | --- | --- | --- |
| Office BPM (Visit 2) | 67.6<br>(50.2, 82.0) | 63.6<br>(30.8, 89.1) | 86.2<br>(68.3, 96.1) | 36.8<br>(16.3, 61.6) | 1.9<br>(0.8, 4.2) | 0.5<br>(0.3, 1.0) | 0.731<br>(0.582, 0.864) |
| Home BPM ($\geq 3$ days) | 89.2<br>(74.6, 97.0) | 81.8<br>(48.2, 97.7) | 94.3<br>(80.8, 99.3) | 69.2<br>(38.6, 90.9) | 4.9<br>(1.4, 17.3) | 0.1<br>(0.1, 0.3) | 0.823<br>(0.613, 0.974) |
| Smartwatch<br>(AI-PPG-ACC-HTN) | 78.4<br>(61.8, 90.2) | 90.9<br>(58.7, 99.8) | 96.7<br>(82.8, 99.9) | 55.6<br>(30.8, 78.5) | 8.6<br>(1.3, 56.3) | 0.2<br>(0.1, 0.5) | 0.838<br>(0.639, 0.969) |

### Error analysis

To understand where and why the AI model made mistakes, we analyzed where the model predictions disagreed with the reference standard. We observed that 17/26 (65.4%) of the AI-PPG-ACC-HTN false negatives were also missed by office BPM screening. In contrast, 9/12 (75%) of the AI-PPG false positives met some criteria for hypertension, particularly during nighttime (Supplemental Table 4). These individuals may benefit from appropriate early care and potential treatment. Furthermore, all false positives had both elevated 24-hour SBP and nighttime SBP or higher, suggesting that all these participants were individuals at higher risk.

To investigate potential confounders in the model, we employed a multivariable logistic regression model to predict the likelihood of a classification error. We found no statistical evidence that the AI-PPG-ACC-HTN classifier’s errors were associated with age, sex, BMI, SBP/DBP levels, or skin tone (Supplemental Table 5).

### Sensitivity analysis for different BP categories

While our primary definition of hypertension was based on 24-hour SBP/DBP ≥130/80, we also evaluated the performance of AI-PPG-ACC-HTN in detecting different categories of elevated BP (24-hour SBP/DBP ≥115/75, ≥125/75, ≥145/90 mmHg) using the same operating point (Supplemental Table 6). AI-PPG-ACC-HTN achieved a sensitivity of 39.9% (95% CI, 32.1%-48.1%), 52.8% (95% CI, 42.9%-62.6%), and 89.5% (95% CI, 66.9%-98.7%) for detecting 24-hour SBP/DBP ≥115/75, ≥125/75, and ≥145/90 mmHg, respectively. Sensitivity increased with increasing BP levels, indicating that AI-PPG-ACC-HTN is more likely to identify individuals with higher BP levels. The sensitivity of AI-PPG-ACC-HTN for detecting sustained hypertension (defined as office SBP/DBP ≥140/90 and 24h SBP/DBP ≥130/90 and daytime SBP/DBP ≥135/85 and night-time SBP/DBP ≥120/70 mmHg) was 100% (95% CI, 69.2%-100.0%). Conversely, the specificity for sustained non-elevated BP (Office SBP <120 and DBP <80, and 24h SBP <115 and DBP <65, and Daytime SBP <120 and DBP <80, and Nighttime SBP <100 and DBP <65 mmHg) was 100% (95% CI, 54.1%-100.0%), demonstrating that all participants classified as positive for hypertension by AI-PPG-ACC-HTN exhibited some signs of elevated BP (Supplemental Table 7).

### AI-PPG-ACC-HTN score is correlated with mean BP levels

Beyond binary classification, the AI-PPG-ACC-HTN score (i.e. the predicted probability of hypertension) was strongly correlated (r=0.66, p<0.001) with mean 24-hour SBP and mean 24-hour DBP (0.63, p<0.001) from ABPM (Supplemental Figure 2). Strong correlations were also observed between office SBP versus mean 24-hour SBP (r=0.70, p<0.001), office DBP versus mean 24-hour DBP (r=0.66, p<0.001), home SBP versus mean 24-hour SBP (r=0.74, p<0.001), and home DBP versus mean 24-hour DBP (r=0.69, p<0.001). The AI-PPG-ACC-HTN score was weakly correlated (r=0.17, p<0.05) with mean heart rate (HR) from 24-hour ABPM. Similarly, weak correlations were also observed between office DBP versus mean 24-hour HR (r=0.20, p<0.005), and between home DBP versus mean 24-hour HR (r=0.19, p<0.01). Neither office SBP nor home SBP were significantly correlated with mean 24-hour HR.

Next, we conducted an analysis to understand how the duration of smartwatch monitoring affected performance. The area under the receiver operating curve (AUROC) increased with more cumulative minutes of sensor data (randomly sampled 2-minute segments) and started to plateau after around 60 minutes of data per day. Performance increased further as the number of observation days increased (Figure 2B). These results support our hypothesis that aggregating many measurements over multiple days of observations can mitigate the errors of individual measurements and challenges of transient confounders.

**Figure 2.**
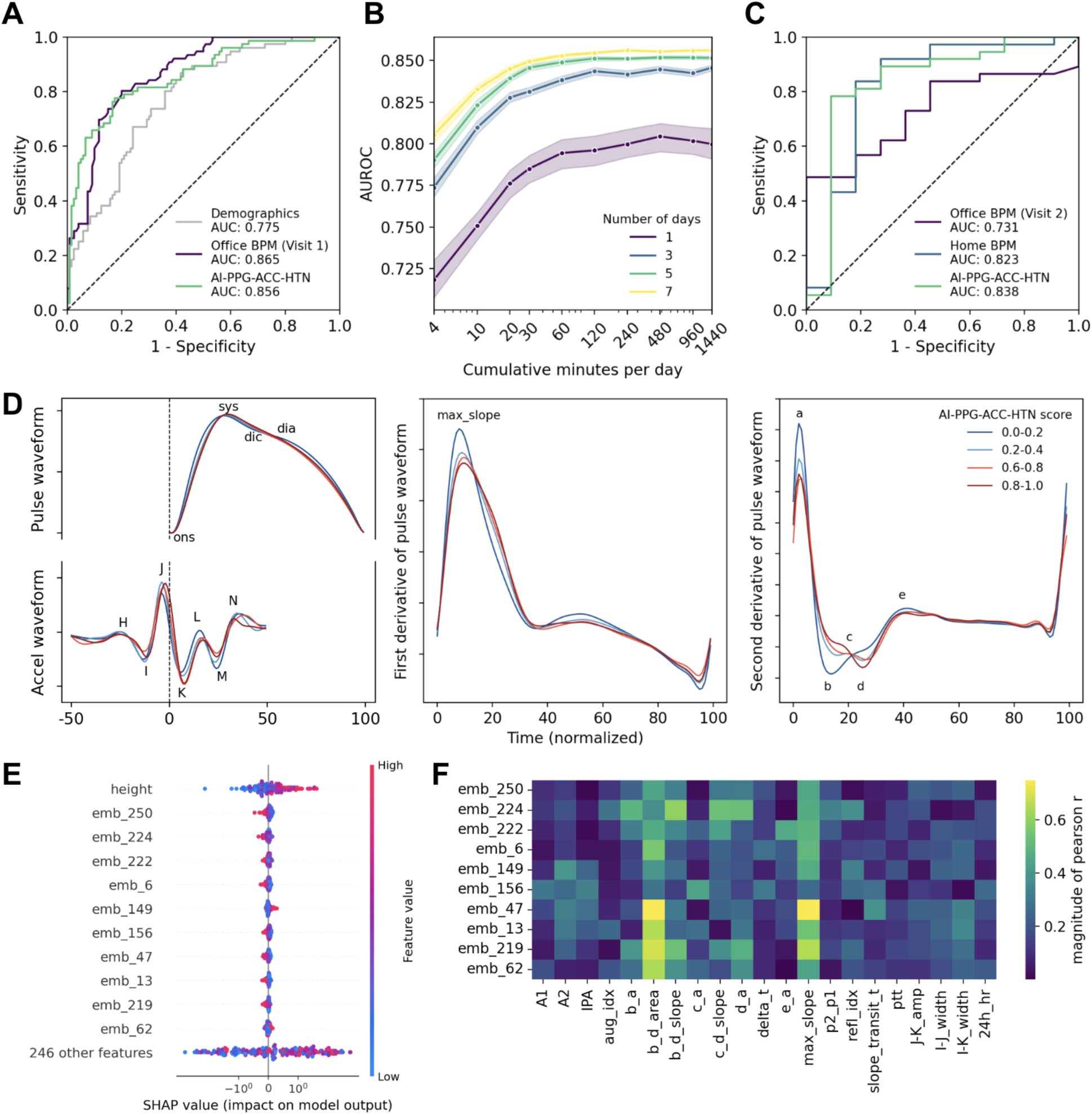
AI-PPG-ACC-HTN performance, explainability analysis, and associations with incident morbidity and mortality. (A) Receiver operating characteristic (ROC) curves showing the performance of a baseline demographics-based model (light gray curve), mean arterial pressure of screening office blood pressure monitoring (office BPM; black curve) and AI-PPG-ACC-HTN (green curve) to identify hypertension. (B) AI-PPG-ACC-HTN performance as a function of wear time. Shaded areas indicate 95% confidence intervals for corresponding colored lines. (C) Performance of repeat office BPM (black curve), home BPM (blue curve), and smartwatch (green curve) to confirm hypertension in participants who initially screened positive for hypertension. (D) Mean pulse waveforms (top left panel), corresponding accelerometer waveforms showing the ballistocardiogram (BCG, bottom left panel), first (middle panel) and second derivatives (right panel) of pulse waveforms from four of the most extreme quintiles of AI-PPG-ACC-HTN scores. Key fiducial points are indicated including the pulse onset (ons), peak (sys), dicrotic notch (dic), diastolic peak (dia) in the PPG waveform, H, I, J, K, L, M, N waves in the BCG waveform, and a, b, c, d, e waves in the second derivative of the PPG waveform. (E) Beeswarm plot of the top features by their Shapley additive explanations (SHAP) values. Lower feature values are colored in blue while higher values are colored in red. (F) Correlation matrix between the top 10 embedding features and hand-engineered PPG and BCG features. Correlations are colored by the magnitude of the Pearson correlation coefficients. A1: area between pulse onset to dicrotic notch; A2: area between dicrotic notch and pulse end; IPA: inflection point area (A2/A1); aug_idx: augmentation index; b_a: b/a ratio; b_d_area: area between b and d wave; b_d_slope: slope of line between b and d, normalized by a; c_a: c/a ratio; c_d_slope: slope of line between c and d, normalized by a; d_a: d/a ratio; delta_t: time between systolic and diastolic peak; e_a: e/a ratio; max_slope: maximum slope during systole; p2_p1: ratio of p2 to p1; refl_idx: reflection index; slope_transit_t: slope transit time; ptt: BCG-PPG pulse transit time; J-K_amp: amplitude between J and K wave; time between J and K wave: time between I and J wave; I-K_width: time between I and K wave; 24h_hr: mean 24-hour heart rate.

### Model interpretability

To gain understanding on what the AI model is paying attention to, we visualized the mean participant-aggregated PPG waveforms from four of the most extreme quintiles of AI-PPG-ACC-HTN scores from lowest to highest probability of hypertension to explore the waveform morphologies associated with model predictions (Figure 2D). Qualitative comparison of the mean PPG waveforms demonstrates that the differences are subtle. PPG waveforms associated with higher AI-PPG-ACC-HTN scores appear to have a slightly reduced pulse width, indicating a smaller change in blood volume, and may suggest stiffer arteries that are less able to expand. Visual changes were more prominent in the first and second derivatives of the PPG waveform. A lower maximum slope of the systolic upstroke and lower d/a ratio were both accompanied by an increasing AI-PPG-ACC-HTN score. The d/a ratio quantifies the relative amplitude of the reflected wave (d) to the initial forward-traveling wave (a). The mean accelerometer waveforms revealed that the BCG signal was captured by the smartwatch. BCG signals associated with higher AI-PPG-ACC-HTN scores appear to have deeper K waves and wider IJK complexes. This pattern appeared similar across age groups (Supplemental Figure 3).

We also analyzed the predictive contributions of the various elements in the waveform embeddings to the hypertension classifier and identified the most important AI-learned features (Figure 2E). We found that these top AI-learned features were strongly correlated with the maximum slope of during systole (r=0.75, p<0.001), the b-d area (r=-0.74, p<0.001), b-d slope (r=0.64, p<0.001) and moderately correlated with the d/a ratio (r=0.52, p<0.001) of the PPG waveform (Figure 2F); these hand-engineered features were consistent with the qualitative visual differences. The top AI-learned features were weakly-to-moderately correlated with hand-engineered features of the BCG waveform such as the I-K width (r=-0.35, p<0.001). This suggests that while the ACC signal is an important component for the model’s performance, its contribution may be more complex than capturing hand-engineered BCG-derived features, potentially also acting as a sophisticated filter for motion or capturing other aspects of mechanical cardiac function not fully represented by traditional BCG features. Many of the hand-engineered features that were correlated to the AI-learned features are known to be associated with BP^23^, signifying that the AI model learned to extract physiologically meaningful features.

## Discussion

In this study, we demonstrate that a widely available consumer smartwatch, without requiring any cuff-based calibration, can detect hypertension with an accuracy comparable to established clinical methods. In a prospective, multicenter study using gold standard 24-hour ABPM as the reference, the AI-PPG-ACC-HTN system detected hypertension with a performance comparable to multi-day home monitoring and with greater sensitivity than single-session office measurements, the status quo for hypertension screening. These findings present a viable strategy for leveraging consumer technology to address the critical global health challenge of undiagnosed hypertension.

The major hurdles for cuffless BP devices may be due in part to noisy cuff BP measurements as labels, but more so from the susceptibility of the PPG waveform to numerous confounding factors including temperature, contact pressure, wrist posture, and motion artifacts. Our approach circumvents these challenges by reframing the objective. Instead of estimating absolute systolic and diastolic pressures at a single moment, we determine an individual’s underlying hypertensive state over time. As these confounding effects are often transient in nature, we reasoned that aggregating many measurements over multiple days of observations would help mitigate isolated effects from individual factors. By leveraging self-supervised learning on a large, unlabeled dataset, our AI system learned to extract physiologically-rich features from PPG and ACC signals. The subsequent aggregation of these features over days allows the persistent signature of hypertension to be distinguished from ephemeral noise. In principle, this is similar to how averaging the many BP measurements over the course of 24 hours helps to mitigate the fact that oscillometric cuff BP devices also yield some individual measurements with appreciable error^19^. As our results indicate, the more readings averaged to create a summary embedding for an individual, the greater the reliability of our approach. While this approach does not produce point-in-time BP estimates, it focuses on the more clinically actionable goal for screening: determining if an individual’s baseline BP is persistently elevated.

Many researchers have relied on the publicly-available MIMIC III waveform database^24^ to evaluate PPG-based algorithms for estimating BP or classifying hypertension. However, the MIMIC dataset comprises patients in the intensive care unit, with many potential confounding effects such as administration of medications and fluids, and is not representative of the general population. Moreover, a recent review revealed that a significant portion of prior papers contained one or more common pitfalls such as data leakage and unreasonable calibration^25^. Our research builds upon and is distinguished from other significant efforts in cuffless hypertension detection, notably the work of Lin et al.^14,15^ and Sola et al.^15^. While these two studies also leverage PPG signals for a classification task, they differ in terms of clinical validation and methodological rigor. The work by Lin et al. demonstrated the feasibility of longitudinal screening, but was validated against home BPM and used cross validation, lacking external validation. The study by Sola et al. also explored high BP detection, but their hypertension label was based on single, intermittent home BPM readings. Critically, their reported performance was aided by a ±8 mmHg exclusion zone. This methodological choice removed borderline cases and simplified the classification task. Furthermore, their data, while extensive, was a retrospective analysis of a self-selected population of commercial device users performing a structured, at-rest task, which differs from a prospective trial in a controlled, diverse cohort under truly free-living conditions. A more direct comparator is Apple’s recently FDA-cleared Hypertension Notification Feature (HTNF) which uses PPG-only features to detect hypertension over a 30-day period; accelerometer data was used only to determine whether the user is sitting still^26^. HTNF achieved a sensitivity of 41.2% and specificity of 92.3% for detecting home BPM ≥ 130/80 mmHg and a sensitivity of 53.7% for detecting home BPM ≥ 135/85 mmHg^16,26^. In contrast, the 65.8% sensitivity achieved by our AI-PPG-ACC-HTN system against ABPM ≥ 130/80 mmHg using a shorter observation period of around 7 days suggests our multimodal approach may offer improvements in detecting at-risk individuals who would otherwise be missed.

Our interpretability analysis shows that the AI features most predictive of hypertension are strongly correlated with established, hand-engineered biomarkers of arterial stiffness and pulse wave reflection associated with BP. This provides evidence that the system is learning genuine vascular pathophysiology reflected in the pulse wave contour, rather than relying on spurious, non-physiological correlations. Changes in the K wave of the BCG, which is caused by deceleration of blood flow in the descending aorta as it is slowed by the peripheral resistance, have been associated with increasing peripheral resistance and arterial inelasticity as in hypertension^27^. The modest contribution from BCG-based features may reflect the challenges of accurately measuring these timings from the wrist in free-living conditions. The strength of our model therefore lies in its ability to synthesize information from a multitude of subtle morphological changes across these waveforms, a task for which AI is well-suited. The robustness of the underlying self-supervised pre-training methodology is further supported by the separate, exploratory analysis using a unimodal version of our encoder on the UK Biobank cohort, demonstrating that the AI-learned PPG features were strongly prognostic for incident hypertension (HR 1.83) and long-term adverse cardiovascular outcomes (Supplemental Information).

Strengths of this study include the prospective, multicenter validation that minimizes selection bias and enhances the generalizability of our findings beyond a single institution. Another strength of the current study is the intentional recruitment of a diverse cohort varied in age, sex, race, and representing a wide range of skin tones. This addresses the critical issue of performance equity for optical-based health sensors. Crucially, we benchmarked our system against 24-hour ABPM, the accepted clinical gold standard for hypertension diagnosis, providing a more robust and clinically relevant assessment than comparisons to less reliable office BPM measurements alone. Moreover, the deliberate exclusion of individuals on antihypertensive medications allowed isolation of the physiological signatures of untreated hypertension, ensuring our model learned fundamental vascular pathophysiology rather than potential confounding effects of medications. Finally, by conducting the validation under free-living conditions and including direct comparisons to both office BPM and home BPM, we have demonstrated the system’s utility in the real-world context for screening, providing evidence for its potential role in the existing clinical landscape.

The performance of any screening tool should be evaluated in the context of its intended use. A positive test result from the smartwatch should act as a trigger for the user to seek formal clinical assessment, including confirmatory measurements according to guidelines with validated cuff-based devices like ABPM or home BPM. While the AI system’s sensitivity of 65.8% is moderate, this figure was higher than initial office BP-based screening (55.3%) conducted in a head-to-head comparison. We note that the sensitivity and specificity of the initial office BP and confirmatory home BP observed in our study were comparable to the previously reported pooled estimates^28,29^ and their 95% CIs substantially overlapped, indicating that they were performed according to best practices. This is particularly significant given that office BP remains the gateway to hypertension diagnosis for most of the population, despite its known limitations, such as susceptibility to the white-coat effect and its inability to detect masked hypertension as well as limitations in access to care in the US. Importantly, the system achieved a high specificity of 90.0% and high PPV of 80.6%, which is important for minimizing the rate of false positives and avoiding undue burden on healthcare resources in a population-scale screening scenario. Furthermore, our error analysis revealed that the majority of false positives from AI-PPG-ACC-HTN had elevated BP by other criteria. This suggests that the AI system’s true clinical specificity is likely higher, as these classifications were not spurious errors but correct identifications of at-risk individuals. Conversely, a large proportion of false negatives were also missed by traditional cuff-based methods, underscoring the challenge of detecting all forms of hypertension with any single modality^30^. One advantage of consumer wearables is that screening can be passively repeated, thus providing multiple opportunities to detect signs of hypertension.

This study has several limitations. We observed a relatively large amount of missingness in data. This attrition was driven primarily by failure of the reference standard (24-hour ABPM) rather than smartwatch data loss. Many participants did not wear the ABPM to sleep and for others, their ABPM files were empty due to participant or study site staff error in accidentally starting a new session after completing the 24 hour wear period or during data upload, which led to the original file being overwritten. This underscores known limitations of cuff-based monitoring, where issues with cuff fit and comfort can lead to lower adherence. However, we observed no significant differences in the demographics and baseline BP of the full enrolled cohort versus the final analysis cohort. Importantly, comparisons to cuff-based BP monitoring devices were performed in a head-to-head manner in the same cohort. Our validation cohort was intentionally selected to exclude individuals with overt cardiovascular disease or those on antihypertensive medications to establish the AI-PPG-ACC-HTN performance in a primary screening population. Its utility for monitoring treatment efficacy or managing BP in patients with established hypertension or those receiving antihypertensive medications for other clinical indications requires further investigation. While our cohort was diverse, validation in larger, more globally representative populations is an important next step to confirm generalizability. Importantly, there is also a need for randomized controlled trials to demonstrate the effectiveness of the proposed AI system in identifying more patients who would have been undiagnosed without AI, and ultimately, to demonstrate improved patient outcomes.

In this paper, we described the development and validation for a new approach to hypertension detection. With hundreds of millions of smartwatches already in use globally, the proposed approach offers a scalable and accessible method to passively and opportunistically screen for hypertension. This could be particularly transformative for reaching the more than 50% of adults with uncontrolled hypertension who are unaware of their condition, a group that is disproportionately younger and has fewer healthcare interactions. By flagging at-risk individuals for follow-up clinical evaluation, this technology could serve as a powerful tool to close the awareness gap, facilitate earlier diagnosis and intervention, and ultimately reduce the long-term incidence of cardiovascular events.

## Methods

### Study Details

All study protocols were approved by an Institutional Review Board (Quorum now known as Advarra, Advarra, Columbia MD and WCG, Puyallup WA). We obtained informed consent from all participants, and the study was conducted in accordance with the principles of the Declaration of Helsinki.

### Skin Tone Measurements

Skin tone was objectively measured using a portable handheld colorimeter (Pantone RM200QC, Pantone LLC, Carlstadt, NJ) that was placed against participants’ wrist. We computed the Individual Typology Angle (ITA°) for each participant and classified them into light, medium and dark skin tone groups following work from Del Bino and Bernard^31^.

### Office BP Measurements

During the initial visit, a set of baseline office blood pressure measurements was taken using the study site’s standard automatic blood pressure devices, following the International Society of Hypertension (ISH) guidelines:

#### Preparation for BP measurements

Participants were instructed to 1) remain still, 2) avoid smoking, caffeinated beverages, or exercise within 30 min before BP measurements, 3) ensure at least five minutes of seated, quiet rest before BP measurements, and 4) sit with back supported and feet on floor, 5) avoid talking.

#### Determining if arm BP differences are present

One BP reading was obtained from each arm, with a two minute interval between readings. Either arm could be measured first. All subsequent BP readings from office, home BPM or ABPM were obtained from the arm with the higher BP value.

#### Obtain 2x further BP readings from the selected arm

A further two BP readings were obtained from the arm selected above. The average of these two BP readings were used as the Office BPM value from Visit 1. During the second (return) visit, a further two BP readings were obtained from the arm selected above, using the preparation as noted above. The average of these 2 BP readings was used as the Visit 2 office BPM value.

### Ambulatory BP Monitoring

Participants were outfitted with an FDA-cleared ABPM (ABPM50, Contec Medical, China), trained on proper use of the device, and instructed to wear the ABPM for 24 hours, starting from the end of the first office visit. The ABPM was configured to measure BP every 20 minutes, from 7 AM to 11 PM, and hourly from 11 PM to 7 AM. Artifactual measurements were removed using thresholds for automatic editing of ABPM data^32^. To minimize participant loss, we considered participants with ≥8 daytime and ≥4 night-time valid readings to have complete ABPM data based on findings that this number of measurements are sufficient to estimate the ambulatory BP levels without meaningful loss of information in hypertension categorization or risk stratification^33^. Daytime period was defined as 9 AM to 9 PM and night-time from 1 AM to 6 AM following recommendations^34^. Mean 24-hour BP was defined as the averages of hourly SBP and DBP mean values (time-weighted averaging)^35,36^.

### Home BP Monitoring

Participants were provided with an automated home BP monitor (Welch Allyn 1700, Welch Allyn Inc., Auburn, NY, USA), with the correct cuff size, and trained on the proper use of the device. On the 2nd through 7th days of the free-living portion of the study, participants were instructed to take at least two sets of readings: one set in the morning after rising and before taking any regular morning medications, and one set in evening before bedtime. Each set of readings consisted of two separate blood pressure measurements using the home BPM, with each home BPM measurement separated by approximately two minutes in between. Instructions on home BPM procedures followed AHA/ACC guidelines, in which participants were instructed to 1) remain still, 2) avoid smoking, caffeinated beverages, or exercise within 30 min before BP measurements, 3) ensure at least five minutes of seated, quiet rest before home BP measurements, 4) sit with back supported and feet on floor, and 5) avoid talking. We considered participants with a minimum of 3 days of home BP measurements to have complete home BP data based on AHA guidelines^18^. BP on Home BP was defined as the average of all available home BP measurements during the study period.

### Smartwatch

Participants were instructed to wear a Pixel watch 3 (Google LLC, Mountain View, USA) configured to record raw data (e.g. PPG, accelerometry etc.) throughout the entire duration of the at-home study, including during sleep. The consent process explicitly detailed the investigational nature of the smartwatch-based AI system and that it was not a validated medical tool.

## Model Development

### Waveform Encoder

To develop a general purpose DL model capable of extracting meaningful representations (embeddings) from wrist PPG and ACC data that are useful for multiple downstream applications, we leveraged approximately 500,000 hours of consented watch sensor data collected across multiple IRB-approved studies conducted by Fitbit to train a general convolutional neural network (CNN) encoder that produces embeddings from input PPG and ACC recordings in a self-supervised manner based on contrastive learning.

The Waveform Encoder uses an EfficientNet B1 architecture with ∼5.1 million parameters. The model inputs are 15-second segments of concomitant PPG and magnitude ACC waveforms stacked channel-wise. The model produces an embedding of size 256. We trained the Waveform Encoder using a self-supervised learning method based on participant contrastive learning^37,38^. The only labels required to train participant contrastive learning are which PPG and ACC comes from which participant. Participant contrastive learning maps the stacked PPG and ACC segments acquired at different times from a given participant to the same region within a contrastive latent space. This approach relies on the shared underlying biology of different PPG and ACC measurements taken from the same participant to form positive pairs; negative pairs constitute PPG and ACC measurements obtained from different participants. The resulting embeddings are constructed in a manner to ensure that different PPG and ACC segments, which arise from the same participant, are more similar to one another relative to PPG and ACC-embeddings arising from different patients.

### Person-level Aggregation

After waveform embeddings are created for all valid segments, they are mean-aggregated by hour of day across days to create up to 24 hour-of-day embeddings. These hour-of-day embeddings are then mean-aggregated to produce a single user embedding. Lastly, the user’s height is z-score normalized and concatenated to the user embedding, forming a vector of 257 elements representing the final person-level embedding.

### Hypertension Classifier

We trained a logistic regression model for hypertension classification with the development dataset using L2 regularization. The input was a 257-length person-level embedding. The labels were the presence of hypertension defined as mean 24-hour ABPM SBP/DBP ≥ 130/80 mmHg. The trained classifier generates a continuous number between 0 and 1 for hypertension classification. We determined the operating point (threshold) by performing 10-fold stratified cross validation on the development dataset. Predictions across all folds were aggregated and the receiving operating characteristic (ROC) curve was computed. Finally, the threshold that maximized sensitivity at a specificity of 90% was identified. We conducted ablation experiments to evaluate the effectiveness of the accelerometer waveform and height as inputs to the model using 10-fold cross validation on the development dataset.

## Subsampling Experiments

Experiments to understand the relationship between the amount of smartwatch sensor data on the performance of the resulting algorithms also were conducted. To understand the relationship between the amount of smartwatch data on the performance of AI-PPG-ACC-HTN, we randomly subsampled 2-minute sensor recordings at different cumulative durations per day (from 4 minutes up to 24 hours per day) and total number of days (from 1 up to 7 days) and measured performance as the area under the receiver operating curve (AUROC).

## Model Evaluation and Statistical Analysis

Performance metrics of sensitivity, specificity, positive predictive value (PPV), and negative predictive value (NPV) are presented using conservative 2-sided Clopper-Pearson exact 95% CIs. Receiver operating curves (ROCs) were plotted by varying the operating threshold applied to the AI-PPG-ACC-HTN score. For office and home BPM, we computed ROCs and AUCs using the mean arterial pressure (MAP) since it was not possible to vary thresholds across the range of possible values for systolic and diastolic BP pairs. We computed the 95% CIs for AUROC via bootstrapping.

To investigate potential sources of bias in the classifier, we employed a multivariable logistic regression model to predict the likelihood of a classification error (N = 196 observations) using the statsmodels package (v0.14.0) in Python. The model included subject age, 24-hour systolic BP, 24-hour diastolic BP, BMI, objectively measured skin tone (ITA°), and sex as independent predictors.

## Feature Importance

To analyze the predictive contributions of the 257 input features (256 embedding elements plus normalized height) to the final logistic regression classifier, we used SHAP (Shapley Additive Explanations)^39^. To properly account for the data distribution and potential feature correlations, the SHAP values were estimated using a reference distribution derived from the training dataset. We then computed these values for all participants in the test set. To determine the global importance of each feature, we calculated the mean absolute SHAP value across all test participants. This metric quantifies the average marginal impact of each feature on the model’s output probability. The features were subsequently ranked by this global importance value, and the top-ranking elements are visualized in a beeswarm plot (Supplemental Figure 4) to illustrate their predictive power and the direction of their effect.

## Acknowledgements

We thank Jennifer Block, Anu Gabbita, Sushalini Sunku, Keerthana Natarajan, Alexandros Pantelopoulos, Tuan Phan, and Tracy Giest for their support, insights and feedback that enabled this research.

## Data Availability

Demographic information, mean BP readings across all modalities (office BPM, home BPM, ABPM) and AI-PPG-ACC-HTN scores and predictions for each participant from the prospective clinical study are made available. This research has been conducted using the UK Biobank Resource under Application Number 65275. Please visit the UK Biobank website, https://www.ukbiobank.ac.uk/, for application procedures. The AI-PPG-HTN scores for the UK Biobank PPG data will be returned to and made available via the UK Biobank.

## Code Availability

A pseudocode implementation of the algorithms is available in Supplementary Information.

## Supplementary Information

### Exploratory incidence analysis in a large population cohort

To explore the prognostic value of the underlying Waveform Encoder features, we conducted a separate, exploratory analysis in a large, population-based cohort (UK Biobank) containing an external set of PPG waveforms. Our analysis included 134,385 PPGs collected at enrollment from as many UK Biobank participants without known clinical history of hypertension and cardiovascular disease (based on ICD codes and self-reports). PPGs from the UK Biobank presented several differences compared to the smartwatch biosignals used to train the Waveform Encoder: firstly, UK Biobank did not record a concomitant accelerometer signal, preventing the use of a multimodal encoder; secondly, UK Biobank waveforms were collected via a finger pulse-oximeter rather than from wrist-worn smartwatch, impacting PPG morphology; finally, the UK Biobank provides just a representative average single beat PPG rather than a longer raw waveform recording, requiring input adaptation. Due to the above differences, we developed a unimodal version of the Waveform Encoder trained on (smartwatch) PPGs only to infer embeddings from the UK Biobank PPGs. To accommodate the encoder’s fixed 15-s input window, normalized UK Biobank single beat PPGs were temporally resampled based on heart rate and sequentially tiled. The resulting waveforms were truncated at the 15-s boundary to ensure exact duration matching. 256-dimensional Waveform Encoder embeddings were then inferred for all participants, and concatenated to an additional scalar element computed by normalizing participant height. Participant vector embeddings were split into tuning (N=58,117) and holdout (N=76,268) sets based on a geographically-motivated partition of the UK Biobank collection sites (tuning: Birmingham, Bristol, Cheadle, Nottingham, Sheffield, Swansea, Wrexham; validation: Liverpool, Middlesborough, Newcastle, Croydon, Hounslow, Reading). A logistic regression model, AI-PPG-HTN, was trained to classify high BP (i.e., an average SBP/DBP >= 140/90 mmHg) on the tuning split and its binary predictions on the holdout set were used as inputs for Cox Proportional Hazard models of clinical outcomes adjusted by baseline demographic risk factors of age, sex, and body mass index. Outcome definitions were based on first occurrences of relevant ICD codes from the electronic health records, or from entries in self reports and primary care notes mapped to ICD10 by the UK Biobank (Supplemental Table 8).

On the hold-out set of 76,268 individuals from distinct geographical sites without previously known hypertension or cardiovascular disease, we tested associations between the AI-PPG-HTN score and incidence of hypertension diagnosis, as well as its sequelae, adjusting for baseline demographic risk factors (Supplemental Figure 4). AI-PPG-HTN stratified cumulative incidence of hypertension (Supplemental Figure 5), and was significantly associated with risk of incident hypertension (hazard ratio, HR: 1.83 [95% CI, 1.77-1.90]). Among 28,321 participants without baseline hypertension classified as positive for hypertension by AI-PPG-HTN, there were 8,024 incident hypertension diagnoses (10-year cumulative incidence of 25.8% [95% CI, 25.3%-26.3%]) compared to 6,698 diagnoses among 47,947 participants classified as negative for hypertension (cumulative incidence of 11.9% [95% CI, 11.6%-12.2%]). AI-PPG-HTN predictions also stratified cumulative incidence of cardiovascular disease and events (Supplemental Figure 5) and were significantly associated with risk of incident diabetes (HR: 1.37 [95% CI, 1.27-1.49]), coronary artery disease (HR: 1.27 [95% CI, 1.19-1.26]), heart failure (HR: 1.25 [95% CI, 1.17-1.34]), stroke (HR: 1.37 [95% CI, 1.17-1.60]), myocardial infarction (HR: 1.45 [95% CI, 1.27-1.49]) and all-cause mortality (HR: 1.18 [95% CI, 1.09-1.27]). There was no statistically significant association between the AI-PPG-HTN predictions and chronic kidney disease. We further found that higher AI-PPG-HTN scores were associated with an increased risk of morbidity and mortality in a dose responsive manner after adjusting for demographic risk factors (Supplemental Figure 6).

These results, while based on a different, unimodal model, demonstrate that the core physiological features learned by our self-supervised Waveform Encoder are not only associated with hypertension but are also prognostic for long-term incident cardiovascular risk and mortality. This highlights the power of the pre-training approach to capture robust, clinically-relevant physiological signatures.

---

**Supplemental Figure 1.**
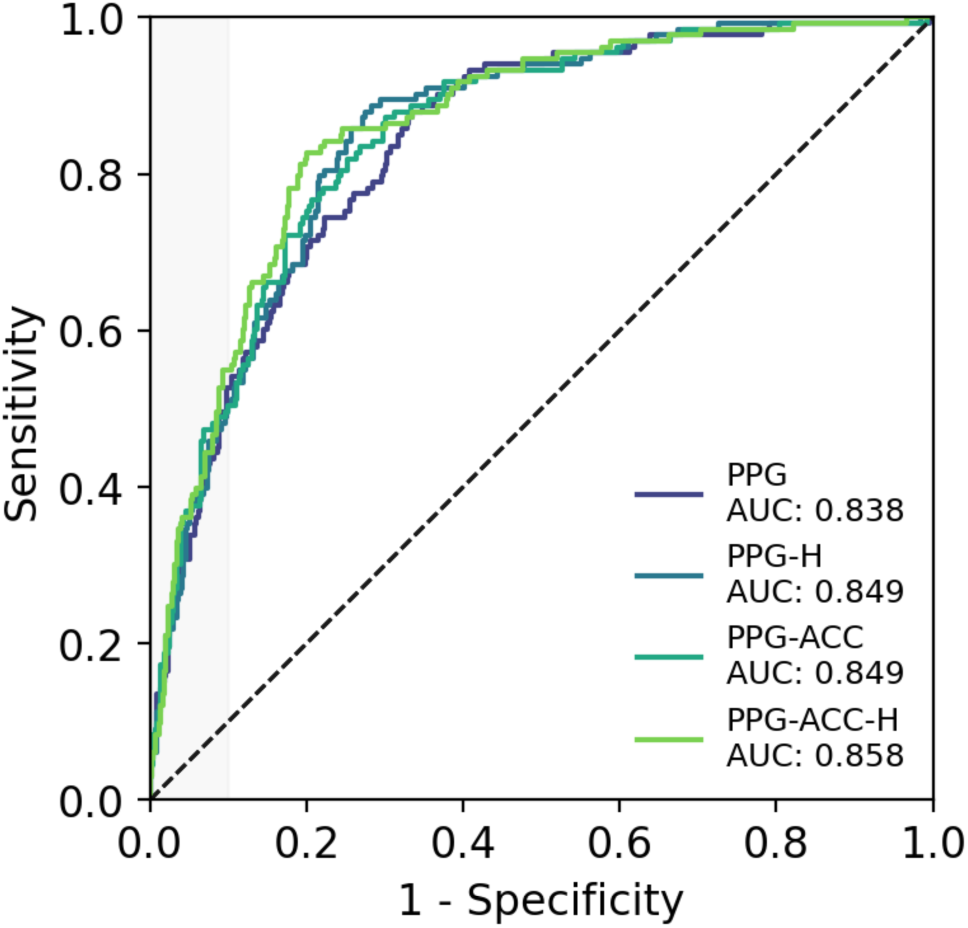
Model Ablations Via Cross-validation on the Development Set. Receiver-operating characteristic (ROC) curves comparing the performance of a photoplethysmography (PPG)-only model (PPG), PPG + height model (PPG-H), PPG + accelerometry (ACC) model (PPG-ACC), and PPG + ACC + height model (PPG-ACC-H).

**Supplemental Figure 2.**
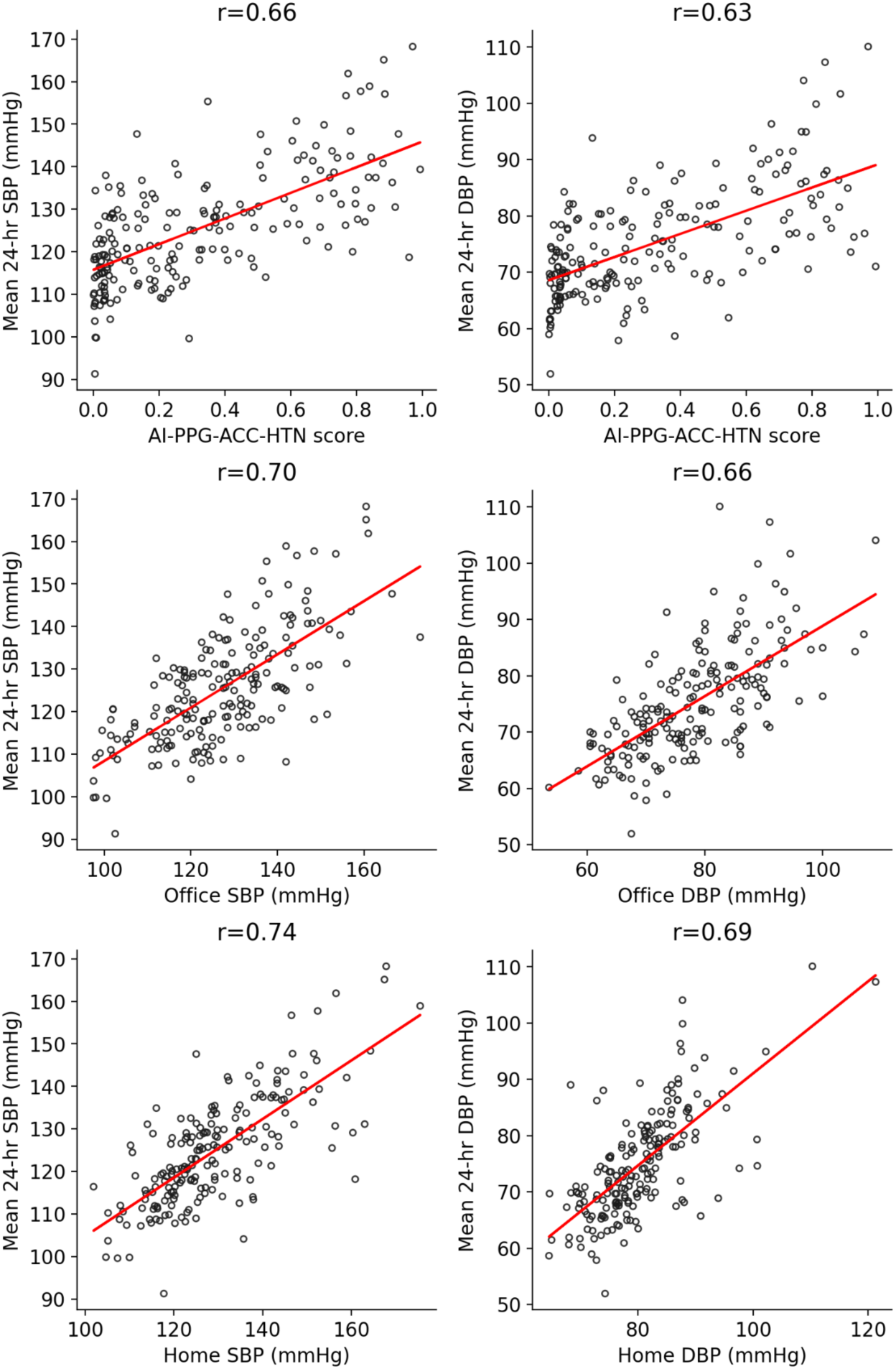
Correlation Between Blood Pressure Monitoring Modalities. Scatterplots of AI-PPG-ACC-HTN score (predicted probability of hypertension), office blood pressure monitoring (BPM) and home BPM versus 24-hour systolic BP (SBP, top) and diastolic BP (DBP, bottom) using ambulatory BPM.

**Supplemental Figure 3.**
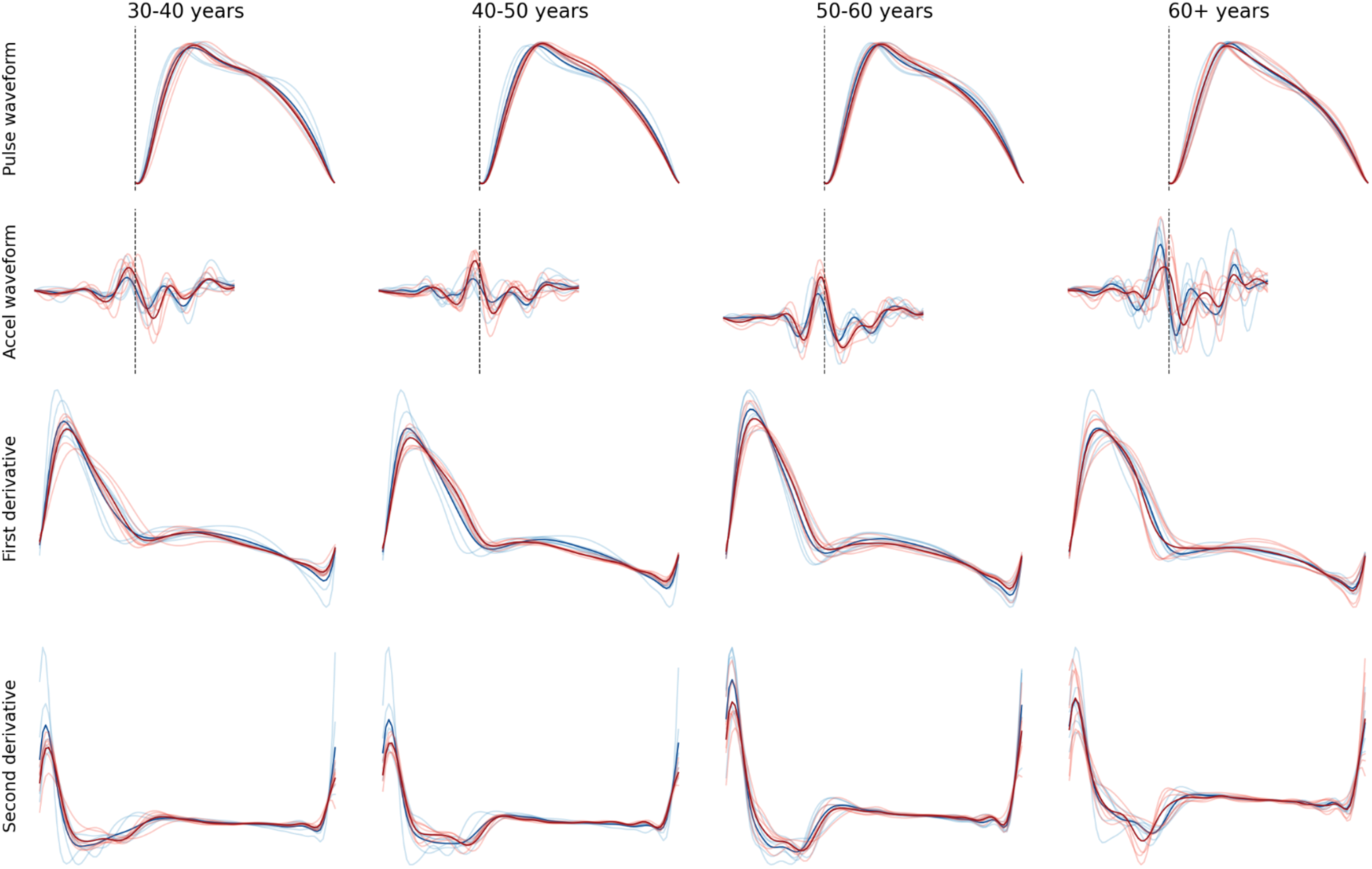
Pulse and accelerometer waveform changes by AI-PPG-ACC-HTN scores and age groups. Mean photoplethysmography (PPG) pulse waveform and corresponding accelerometer waveforms (top row), the first (middle row) and second (bottom row) derivatives of the PPG waveform across different age groups. Waveforms from the highest AI-PPG-ACC-HTN scores are shown in red; those with the lowest hypertension scores are in blue. Both individual-level waveforms (light) and the mean waveform across individuals (bold) are displayed.

**Supplemental Figure 4.**
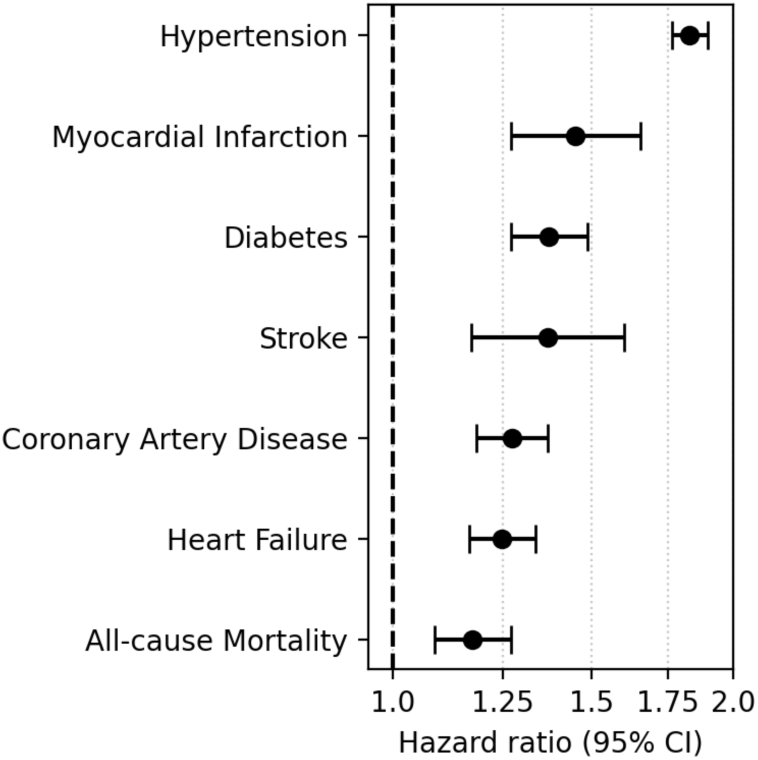
Associations between AI-PPG-HTN and incident cardiovascular outcomes. AI-PPG-HTN classifications were significantly associated with risk of 10-year incident hypertension, diabetes, coronary artery disease, heart failure, stroke), myocardial infarction and all-cause mortality after adjusting for age, sex and body mass index in a population-based cohort (UK Biobank) without baseline hypertension or cardiovascular disease.

**Supplemental Figure 5.**
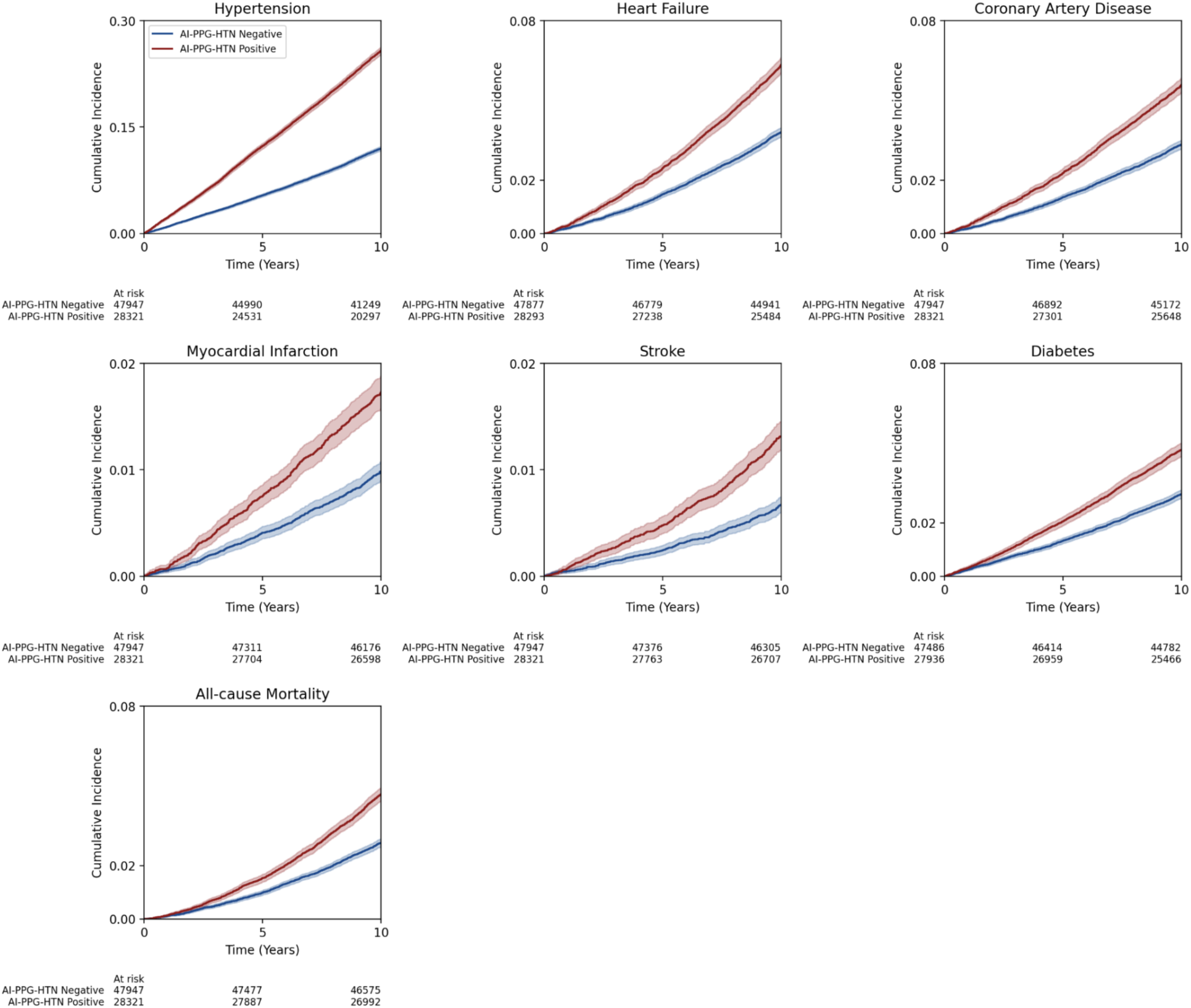
Cumulative incidence of cardiovascular disease and events by AI-PPG-HTN model classifications. Positive classification of hypertension by the AI model was associated with a greater risk of incident hypertension, heart failure, coronary artery disease, myocardial infarction, stroke, diabetes and all-cause mortality. Participants with prevalent cardiovascular disease (previous myocardial infarction, stroke, cardiac arrest, ischemic heart disease, atrial fibrillation, heart failure, or aortic valve disorder) at baseline were excluded. Participants with prevalent diabetes were excluded from analysis of incident diabetes only. p<0.005 for all.

**Supplemental Figure 6.**
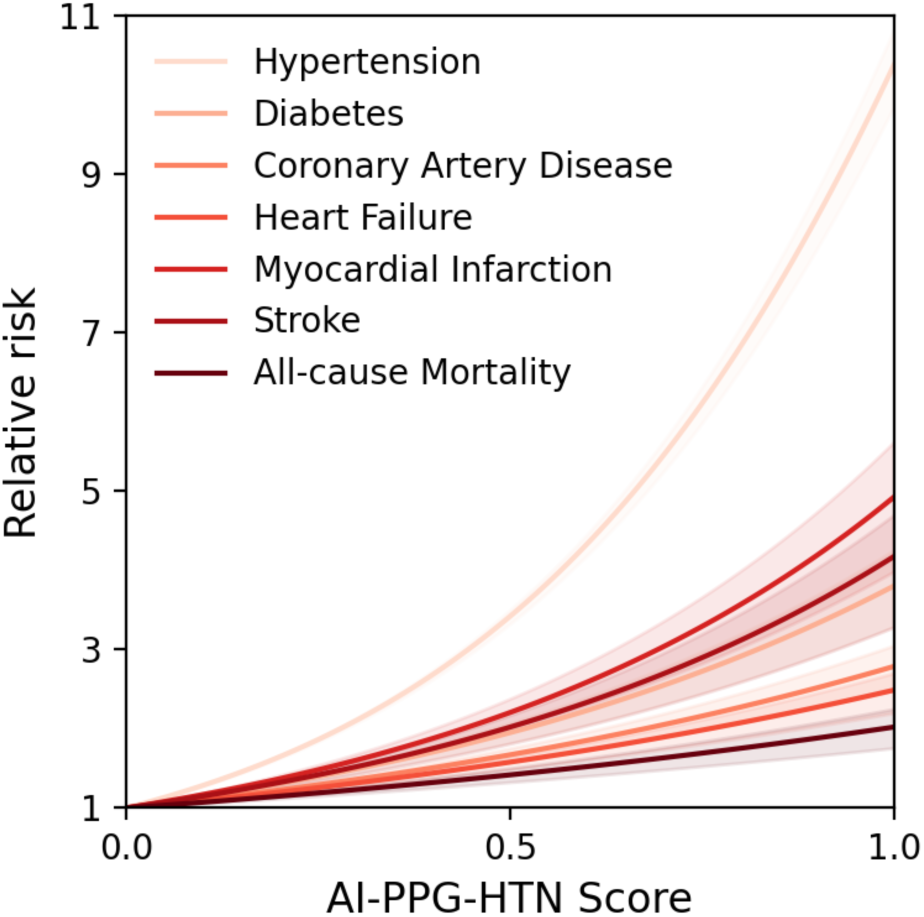
Relative risks of incident cardiovascular disease and events by AI-PPG-HTN score. Higher AI-PPG-HTN scores were associated with an increased risk of hypertension and its sequelae, including diabetes, coronary artery disease, heart failure, myocardial infarction, stroke and all-cause mortality in a nonlinear dose responsive manner after adjusting for age, sex and body mass index. Shaded areas indicate lower and upper quartiles for corresponding colored lines.

**Supplemental Table 1.** Participant Characteristics in Waveform Encoder Dataset.

| Characteristic |  | Training | Tuning | Combined |
| --- | --- | --- | --- | --- |
| Total Participants |  | N = 1,614 | N = 495 | N = 2,109 |
| Age, years |  | 40.5 ± 13.3 | 41.5 ± 14.0 | 40.7 ± 13.5 |
|  | Missing | 13 (0.8%) | 5 (1.0%) | 18 (0.9%) |
| Female |  | 572 (35.7%) | 169 (34.1%) | 741 (35.1%) |
|  | Missing | 13 (0.8%) | 5 (1.0%) | 18 (0.9%) |
| BMI, kg/m<sup>2</sup> |  | 26.5 ± 6.0 | 26.9 ± 5.6 | 26.6 ± 5.9 |
|  | Missing | 13 (0.8%) | 5 (1.0%) | 18 (0.9%) |
| Skintone (Monk skin tone, MST) | MST 1-4 | 644 (39.9%) | 201 (40.6%) | 845 (40.1%) |
|  | MST 5-7 | 319 (19.8%) | 112 (22.6%) | 431 (20.4%) |
|  | MST 8-10 | 21 (1.3%) | 20 (4.0%) | 41 (1.9%) |
|  | Missing | 630 (39.0%) | 162 (32.7%) | 792 (37.6%) |

**Supplemental Table 2.**
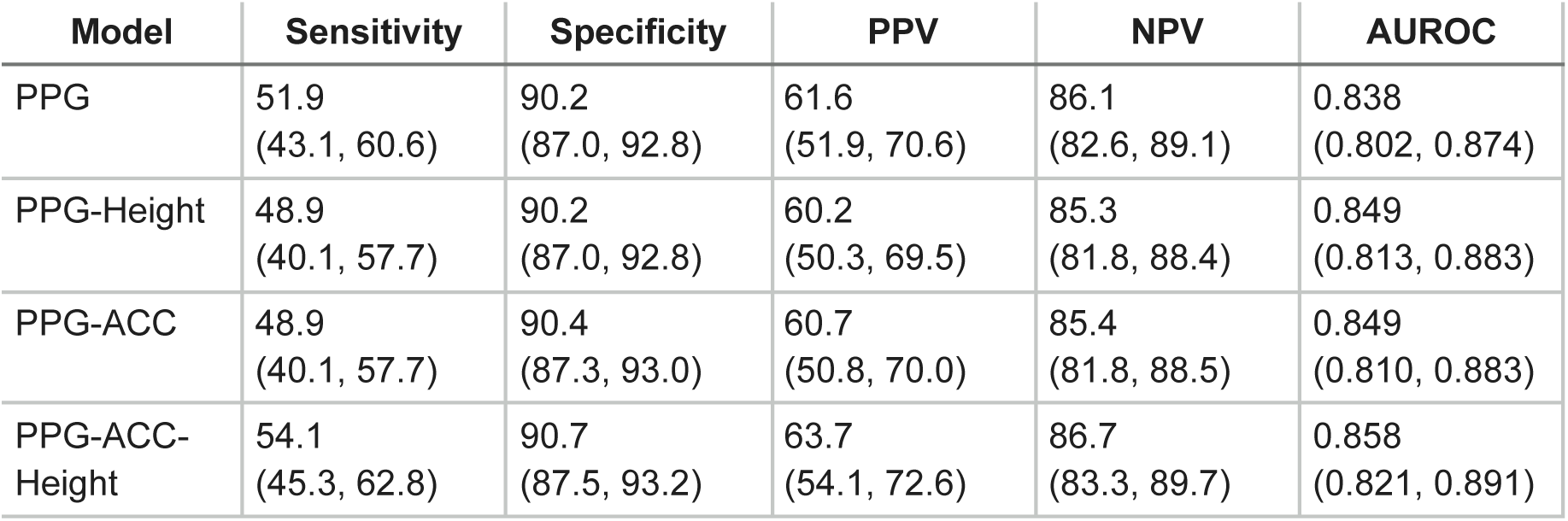
Cross Validation Performance (10-fold) of Various Models to Identify Hypertension in the Development Dataset.

**Supplemental Table 3.**
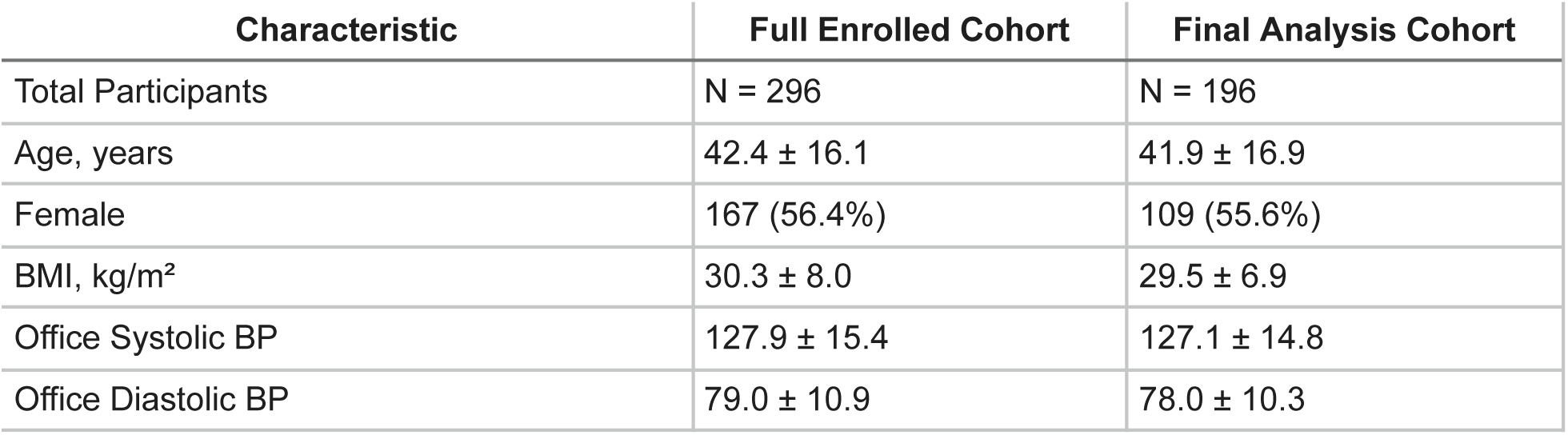
Summary of Participant Characteristics in the Final Analysis versus Incomplete Data Cohort.

**Supplemental Table 4.**
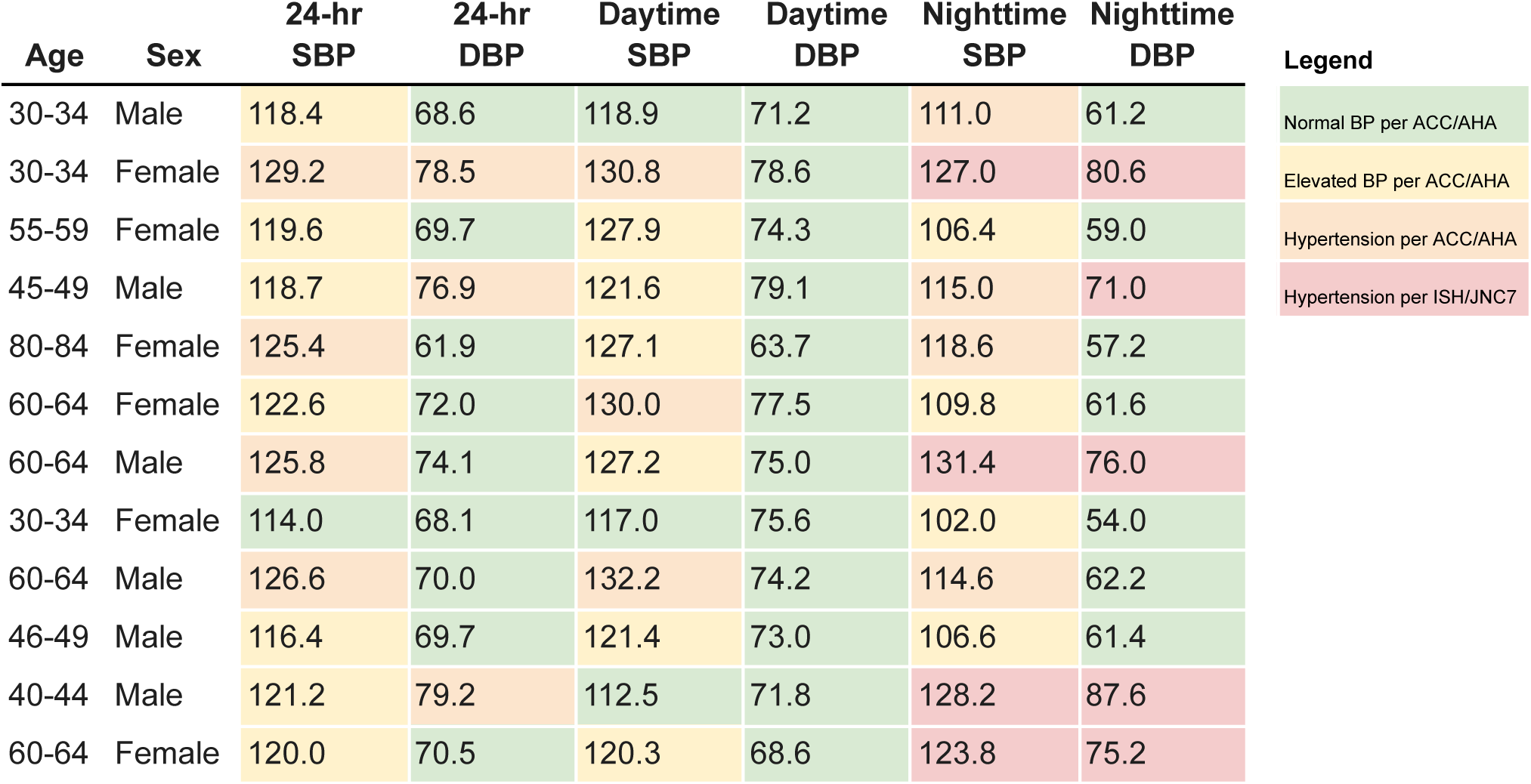
False Positives by AI-PPG-ACC-HTN Based on Hypertension Defined By 24-hour Ambulatory Blood Pressure Monitoring (≥130/80 mmHg)

**Supplemental Table 5.** Multivariate Logistic Regression Results.

| Dep. Variable | pred_error |  | No. Observations: | 196 |  |  |
| --- | --- | --- | --- | --- | --- | --- |
| Model: | Logit |  | Df Residuals: | 189 |  |  |
| Method: | MLE |  | Df Model: | 6 |  |  |
| Log-Likelihood: | -91.864 |  | Pseudo R-squared: | 0.04698 |  |  |
| Converged: | True |  | LL-Null: | -96.392 |  |  |
| Covariance Type: | nonrobust |  | LLR p-value: | 0.1704 |  |  |
|  | coef | std err | z | P>[z] | [0.025 | 0.975] |
| const | -4.9977 | 1.802 | -2.773 | 0.006 | -8.530 | -1.465 |
| 24h_sbp | 0.0235 | 0.907 | 0.907 | 0.364 | -0.027 | 0.074 |
| 24h_dbp | -0.0077 | 0.033 | -0.236 | 0.813 | -0.072 | 0.056 |
| age | 0.0135 | 0.013 | 1.066 | 0.286 | -0.011 | 0.038 |
| bmi | 0.0221 | 0.028 | 0.779 | 0.436 | -0.034 | 0.078 |
| skintone (ITA) | -0.0076 | 0.005 | -1.413 | 0.158 | -0.018 | 0.003 |
| sex_is_male | 0.0263 | 0.417 | 0.063 | 0.950 | -0.791 | 0.843 |

**Supplemental Table 6.** Test Accuracy of AI-PPG-ACC-HTN to Identify Various Levels of High BP.

| **BP Level** | **Sensitivity** | **Specificity** | **PPV** | **NPV** | **AUROC** |
| --- | --- | --- | --- | --- | --- |
| 24h ABPM $\geq 145/90$ | 89.5<br>(66.9, 98.7) | 74.6<br>(67.5, 80.8) | 27.4<br>(16.9, 40.2) | 98.5<br>(94.7, 99.8) | 0.866<br>(0.793, 0.932) |
| 24h ABPM $\geq 125/75$ | 52.8<br>(42.9, 62.6) | 93.3<br>(86.1, 97.5) | 90.3<br>(80.1, 96.4) | 62.7<br>(53.9, 70.9) | 0.852<br>(0.797, 0.901) |
| 24h ABPM $\geq 115/75$ | 39.9<br>(32.1, 48.1) | 97.7<br>(87.7, 99.9) | 98.4<br>(91.3, 100.0) | 31.3<br>(23.6, 39.9) | 0.815<br>(0.742, 0.874) |

**Supplemental Table 7.** Test Accuracy of AI-PPG-ACC-HTN for Sustained Hypertension and Non-Elevated BP.

| **BP Category** | **Sensitivity** | **Specificity** | **PPV** | **NPV** | **AUROC** |
| --- | --- | --- | --- | --- | --- |
| Sustained HTN<br>(Office BPM $\geq 140/90$ and<br>24h ABPM $\geq 130/90$ and<br>Daytime ABPM $\geq 135/85$ and<br>Night-time ABPM $\geq 120/70$) | 100.0<br>(69.2, 100.0) | 72.0<br>(65.0, 78.4) | 16.1<br>(8.0, 27.7) | 100.0<br>(97.3, 100.0) | 0.901<br>(0.848, 0.947) |
| Sustained Non-Elevated BP<br>(Office SBP $< 120$ and DBP<br>$< 80$, and 24h SBP $< 115$ and<br>DBP $< 65$, and Daytime SBP<br>$\geq 120$ and DBP $< 80$, and<br>Nighttime SBP $< 100$ and<br>DBP $< 65$) | 32.6<br>(26.0, 39.8) | 100.0<br>(54.1, 100.0) | 100.0<br>(94.2, 100.0) | 4.5<br>(1.7, 9.5) | 0.885<br>(0.663, 0.990) |

**Supplemental Table 8.**
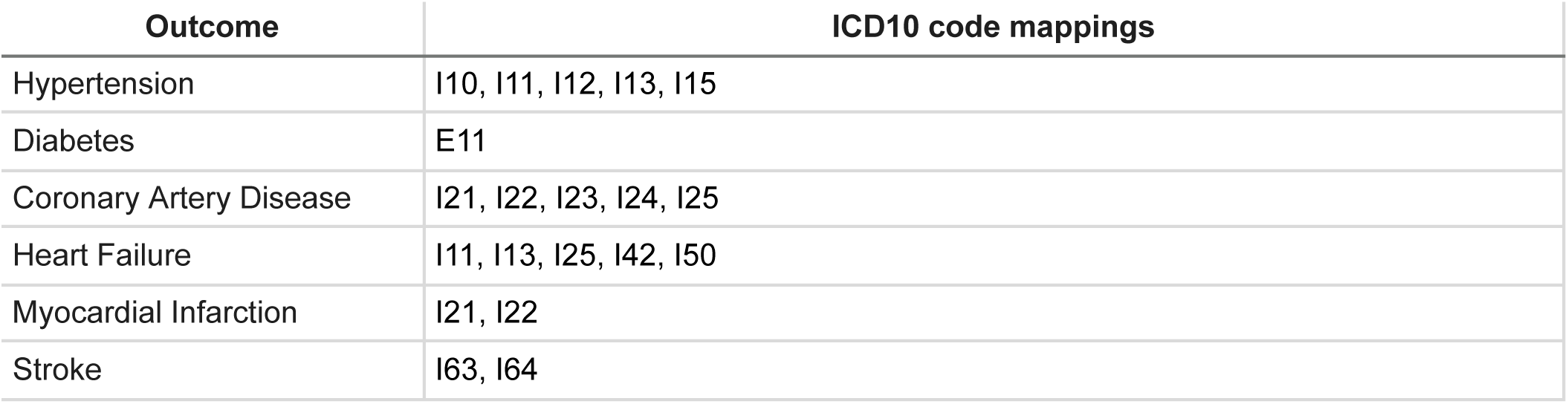
Clinical outcome definitions for the incidence analysis in UK Biobank.

